# Estimating the Impact of Pre-Exposure Prophylaxis (PrEP) on Mortality in COVID-19 Patients: A Causal Inference Approach

**DOI:** 10.1101/2023.03.16.23287365

**Authors:** Ajan Subramanian, Yong Huang, Melissa D. Pinto, Charles A. Downs, Amir M. Rahmani

## Abstract

Traditional machine learning (ML) approaches learn to recognize patterns in the data but fail to go beyond observing associations. Such data-driven methods can lack generalizability when the data is outside the independent and identically distributed (i.i.d) setting. Using causal inference can aid data-driven techniques to go beyond learning spurious associations and frame the data-generating process in a causal lens. We can combine domain expertise and traditional ML techniques to answer causal questions on the data. Hypothetical questions on alternate realities can also be answered with such a framework. In this paper, we estimate the causal effect of Pre-Exposure Prophylaxis (PrEP) on mortality in COVID-19 patients from an observational dataset of over 120,000 patients. With the help of medical experts, we hypothesize a causal graph that identifies the causal and non-causal associations, including the list of potential confounding variables. We use estimation techniques such as linear regression, matching, and machine learning (meta-learners) to estimate the causal effect. On average, our estimates show that taking PrEP can result in a 2.1% decrease in the death rate or a total of around 2,540 patients’ lives saved in the studied population.

## I. INTRODUCTION

Causal inference refers to the process of determining the causal relationship between an effect on some outcome. The goal is to estimate causal effects given observed variables and control for any potential sources of bias that can affect this estimate [1]. There are plenty of fields that can benefit from causal analysis, like medicine, economic policy evaluation, and digital marketing, to name a few. Its power lies in the ability to go beyond identifying spurious correlations in observational data and asking more fundamental questions on how the data is structured and the mechanisms that generate it. Machine learning techniques such as supervised and unsupervised learning often assume that training data is independent and identically distributed (i.i.d). One of the main reasons for the success of these algorithms relies on the fact that they can capture complicated associations in the observed data accurately. However, one major challenge for these methods is that they need to be able to operate effectively when domain shifts occur. Moreover, they fail to account for interventions in the data. Such data-driven methods try to achieve generalizability to unseen examples by training on copious amounts of samples. For example, state-of-the-art convolutional neural networks are trained on millions of images to detect objects accurately. However, generalizing outside the i.i.d regime in the real world, where distributions are constantly altered, requires such algorithms to go beyond identifying mere associations in the data. They need to understand the data-generating mechanisms involved between the variables and capture their relationships in a causal way [2], [3], [4], which can be done through structural causal models (SCM) or models representing causal relationships between variables. Causal Machine Learning (CausalML) has gained much traction in the research community. It is a framework where traditional machine learning techniques can utilize the causal structure of the observed data [5].

The use of causal inference in medicine is driven by estimating the average treatment effect (ATE) of a particular intervention in a population. Specifically, learning the optimal treatment policy for a single patient is one of the promising goals of applying such techniques. The gold standard for estimating such causal effects in a clinical setting is a Randomized Controlled Trial (RCT). However, such data is often expensive, time-consuming to collect, and may sometimes even be unethical, like measuring the effect of smoking on lung cancer. Therefore, there is a need to estimate these causal effects from observational data alone [6]. Causal inference can help us achieve this if we specify the relationship between observed variables in a causal graph. Transformation of observational data into a causal graph requires thorough identification of variables, such as confounders, mediators, and colliders, which have the potential to create non-causal pathways and bias our estimates [7]. This transformation necessitates a strong understanding and expertise in the relevant domain.

Given the ongoing COVID-19 pandemic and its impacts, it is crucial to investigate potential treatment options that could be used alongside existing interventions such as vaccines to minimize the risk of long-term symptoms. A medical study [8] has suggested that PrEP (Pre-Exposure Prophylaxis), which was traditionally used to prevent HIV transmission in at-risk populations [9], [10], could be a promising avenue to explore. With the help of causal inference, it is possible to rigorously and rapidly assess the causal impact of PrEP on specific COVID-19-related outcomes while using observational data. If the results obtained from the causal analysis are satisfactory, it can justify the need for an RCT to further validate this claim.

In this paper, we investigate the causal effect of PrEP on mortality in COVID-19 patients from the University of California COVID Research Dataset (UC-CORDS) [11]. Our contributions are multifold as a case study for applying causality to real-world medical data. Our team of medical experts, utilizing their extensive knowledge and experience, facilitates the initial identification of a set of variables that are pertinent to both the administration of PrEP treatment and mortality. Subsequently, we construct a causal graph that clearly defines the relationship between these observed variables. We then employ a range of estimation algorithms to calculate the individual and average treatment effects for our dataset. The results of our causal analysis provide valuable insights into the efficacy of PrEP in COVID-19-positive patients, allowing us to draw well-informed conclusions.

## II. BACKGROUND

### A. PrEP for COVID-19

According to the United States Department of Health & Human Services, there are currently 38.4 million people across the globe living with HIV, with an estimated 1.5 million individuals acquiring HIV in 2021. In the U.S., approximately 1.2 million people have HIV, with an estimated 13% unaware of the infection, and HIV infections continue to affect racial, ethnic, and sexual minorities disproportionately [12]. Recent efforts to reduce transmission of HIV include highly active antiretroviral therapy (HAART) among those living with HIV and the use of pre-exposure prophylaxis (PrEP) among those at risk.

Data show that the use of HAART and PrEP significantly reduces, if not eliminates, the potential to transmit the virus through sexual activity or injection drug use [9], [10]. Among those with HIV, these medications suppress viral replication. Several HAART regimens are available for those with HIV, and three medication regimens are approved for use as PrEP. With the urgency of the COVID-19 pandemic, data from multiple studies suggested that the presence of chronic conditions increases the likelihood of developing severe COVID-19 disease and death [13]. HIV and being immunocompromised were identified as potential comorbidities that could lead to worse outcomes from COVID-19.

Prior work has speculated that PrEP or HAART could benefit those who contract SARS-CoV-2, which causes COVID-19. Fernandes et al. [8] identified that the use of PrEP was associated with lower self-reporting of COVID-19-related symptoms. However, a study by Ayerdi [14] failed to find a benefit to PrEP in mitigating symptoms. Therefore, the purpose of this study was to identify if the use of HAART or PrEP affected mortality from COVID-19.

### B. Causal Inference Background

The goal of causal inference is to estimate the causal effect of a treatment on an outcome while controlling for certain observed variables that can bias this effect. More specifically, we want to measure treatment X’s average treatment effect (ATE) on outcome Y on a population while adjusting for a set of covariates Z.

The first step in estimating these effects is constructing a Directed Acyclic Graph (DAG) that captures the causal relationships between the observed variables. Construction of this graph requires relevant domain expertise. Once this graph is constructed, the next step is identifying the causal effects. Identifying this effect requires making some untestable assumptions in an observational setting. The most important one is the ignorability assumption which states that there are no unmeasured confounders [1]. Confounders and colliders are two sources of bias in a causal graph and can affect the estimate if not handled carefully. Some of the relevant terms for causal inference are described in detail below:

1. *Confounder:* A confounder is a variable that affects both the treatment and the outcome. For example, age is a confounder for the association between smoking and lung cancer risk because age affects both the probability of smoking and the risk of getting lung cancer. In the figure below, Z is a confounder because it is a common cause of both X and Y. Controlling for Z “blocks” the spurious non-causal path between X and Y. Hence, confounders should be controlled for in any causal analysis.

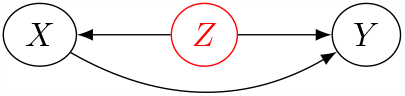
2. *Mediator:* A mediator is a variable through which a treatment causes an outcome. For example, the amount of tar deposited in the lungs as a result of smoking is a mediator of the effect of smoking on lung cancer. In the figure below, Z is a mediator because it mediates the effect of X on Y. Controlling for Z “blocks” the non-causal path between X and Y. Controlling for mediators is feasible if it also blocks confounding variables.

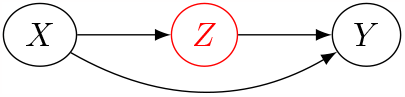
3. *Collider:* A collider is a variable that is caused by at least two other variables. For example, if the quality of life is affected by smoking cessation (treatment) and lung cancer (outcome), this variable would be a collider. Controlling for quality of life will cause a spurious non-causal association between smoking cessation and lung cancer [7]. In the figure below, Z is a collider because it is a child of both X and Y. Controlling for Z “unblocks” the non-causal path between X and Y. Hence, colliders should NOT be controlled for in most cases. Controlling colliders is feasible if their parents are controlled too.

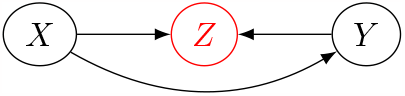
4. *Backdoor Criterion:* To measure the direct effect of X on Y, we have to ensure that we are eliminating these spurious correlations, which is done by ensuring all the non-causal paths from X to Y are blocked off. After identifying these variables, we can apply the “backdoor criterion” which states that “given an ordered pair of variables (X,Y) in a DAG G, a set of variables Z satisfies the backdoor criterion relative to (X,Y) if no node in Z is a descendent of X, and Z blocks every path between X and Y that contains an arrow into X” [15]. Simply put, it keeps the direct causal path from X to Y while blocking off all non-causal spurious paths. If the backdoor criterion is satisfied, we can compute the causal effect of X on Y by the following formula:

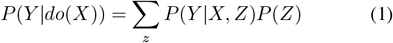
5. *Structural Causal Model:* We can also represent our causal diagrams in the form of a structural causal model (SCM), which describes the state of the variables and how they relate to the distribution of our dataset. Mathematically speaking, an SCM defines the relationship between a set of endogenous or observed variables (V) and exogenous or unobserved variables (U) through a set of functions (F). Each SCM is associated with DAG G where each node in G is a variable in U or V, and each edge is a function in F.
6. *Counterfactuals:* The treatment effect for each patient under study is the difference between two potential outcomes. One outcome is if the unit is exposed to the treatment, and the other is if they are not. The fundamental problem of causal inference is that we can only measure one potential outcome for each patient. As a result, counterfactuals or what would have happened had the patient received the opposite treatment are what causal estimation techniques need to compute. By using a set of assumptions, these estimation techniques can compute the unobservable counterfactual outcome.

## III. METHODS

This section describes how we applied causal inference to our specific case study.

### A. Dataset

The UC-CORDS Dataset [11] consists of comprehensive de-identified health data collected across all facilities in the University of California (UC) Health system. It comprises data from patients admitted to 19 professional schools, five academic medical centers, and 12 hospitals. It contains records of more than 700,000 patients and provides a wide range of information, including medical history, medications, and lab tests of admitted patients. However, in this study, we only considered a limited set of factors that had a direct impact on both taking PrEP medication and mortality.

In order to perform our causal estimation, it was necessary to divide the patients into two distinct and non-overlapping groups, the treatment group and the control group. Both groups consisted of individuals who had tested positive for COVID-19. The treatment group was comprised of patients who had received PrEP medication, while the control group consisted of patients without a history of taking this medication. The corresponding ICD-10 codes for lab tests, demographic information, pre-existing conditions, and medications were used to extract relevant data from our EHR dataset. This resulted in a total of slightly less than 121,000 patients and 1,550 patients within the treatment group alone.

### B. Causal Graph

Our team of medical experts provided assistance in identifying a list of relevant observable variables. Our proposed DAG is presented in Figure 1. Given that the treatment group was primarily composed of white males over the age of 40, it was deemed crucial to include variables such as race, gender, and age due to their potential confounding effects. Additionally, information regarding the individual’s HIV diagnosis, the number of COVID-19 infections, and the number of vaccinations received were also incorporated due to their impact on mortality.

**Fig. 1:**
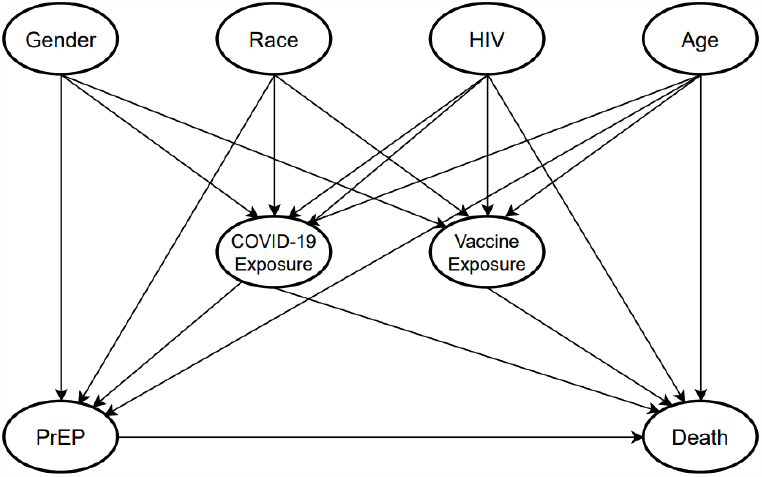
Proposed causal graph with all observed variables

### C. Adjusting for Bias

In our causal graph, the direct confounding variables we need to control for are a patient’s HIV diagnosis and age because both of these variables affect the likelihood of taking PrEP and mortality. Additionally, controlling for colliders like “COVID-19 Exposure” and “Vaccine Exposure” will bias our causal estimate if not handled appropriately and hence should be removed from the analysis. It is important to note that we also need to control gender and race because they directly affect PrEP and indirectly affect death through COVID-19 exposure and vaccine exposure. This is due to the fact that these two exposure variables serve as mediators for both pathways, from race to death and from gender to death. Thus, it is necessary to control for gender and race in order to prevent their direct impact on PrEP and their indirect effect on death through the mediators.

Figure 2 shows the resulting graph after adjusting for these biases. Equation 2 shows the backdoor criterion for our specific problem. Here, PrEP is the treatment, death is the outcome, and Z is the set of all confounders we need to adjust.

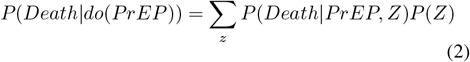

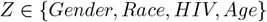

**Fig. 2:**
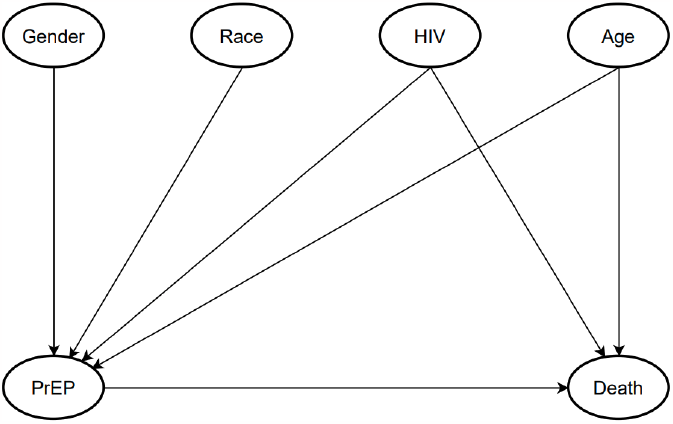
Proposed causal graph after adjusting for bias

### D. Structural Causal Model (SCM)

Given below is the corresponding SCM to our causal graph:

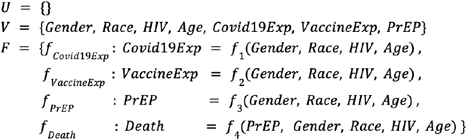

We do not have any exogenous variables (U) that are confounders in our model, so it is represented as an empty set. Since COVID-19 and vaccine exposures are both colliders, there is no need to estimate *f*_1_ and *f*_2_. Moreover, *f*_3_ will be replaced by a single value (0 or 1) when estimating the causal effect. The only function we will need to estimate is *f*_4_.

### E. Causal Estimation

The estimation of causal effects consists of a family of statistical methods. Since we assume we observe all confounders, we can use estimation methods under unconfoundedness, which include matching, reweighting methods like Inverse Probability Weighting (IPW), and meta-learner methods like S-, T-, X-, and R-learners [16], [17], [18].

- Matching: Matching is a technique to pair up similar observations from the treatment group and control group; thus, the unconfounded assumption could be satisfied, and methods such as nearest neighbors could be applied for matching when the appropriate distance metric is defined.
- Inverse Probability Weighting: One downside of the matching method is that it tends only to use some of the data since unmatched observations are discarded. Inverse probability weighting addresses this problem by assigning weights (the probability of receiving treatment)to every observation, and the weights can be generated using methods such as logistic regression.
- Meta-learner: Meta learners deploy off-the-shelf machine learning predictive models for estimation with flexible selections of machine learning models. S-leaner uses a single model to predict treatment effect, while T-learner splits the data and uses one model for each treatment. X-learner builds on top of the T-learner by adding an inverse probability weighting procedure.

## IV. RESULTS AND DISCUSSION

We compute the average treatment effect (ATE), average treatment effect on the control group (ATC) and treatment group (ATT) from five estimators: ordinary least squares regression (OLS), matching, and three meta-learners (S, T, X-Learners). The meta-learners use a light gradient boosting regressor to compute the counterfactual outcomes for each patient.

Table I presents our causal estimates using the above estimators. The first half of the table computes these estimates using all the observed variables, including the colliders. The second half of the table computes them after controlling just the confounders. The numbers in this table are represented as a fraction of the population. For example, an estimate of |0.02| translates to 2% of the studied population.

**TABLE I:** Causal Estimates of taking PrEP on Mortality

| Estimators | OLS | Matching | S-Learner | T-Learner | X-Learner |
| --- | --- | --- | --- | --- | --- |
| All observed variables |  |  |  |  |  |
| ATE | -0.022 | -0.027 | -0.010 | -0.018 | -0.020 |
| ATC | -0.022 | -0.027 | -0.010 | -0.018 | -0.020 |
| ATT | -0.018 | -0.007 | -0.004 | -0.007 | -0.008 |
| Only confounders |  |  |  |  |  |
| ATE | -0.022 | -0.025 | -0.015 | -0.021 | -0.023 |
| ATC | -0.022 | -0.025 | -0.015 | -0.021 | -0.023 |
| ATT | -0.020 | -0.010 | -0.006 | -0.008 | -0.008 |

We can observe that the treatment effects are all negative, meaning there is a reduced mortality risk in taking PrEP medication for COVID-19-positive patients. The ATE scores estimated the difference in treatment effects when everyone in the population received the treatment versus when no one in the population received the treatment. The ATC and ATT scores assess this difference in only the control group and treatment groups, respectively. All three scores for most estimators have a higher magnitude when we only control for confounders. Controlling for all observable variables, including colliders, underestimates the actual causal estimate. Across all estimators, an analysis of the ATE scores estimated from considering only confounders demonstrates a 2.1% decrease in the mortality rate when PrEP is taken as a treatment. This equates to approximately 2,540 patients, when applied to a substantial patient population of 121,000 individuals in our study. Furthermore, when averaging the ATE scores estimated from all observable variables, the results indicate a 1.9% decrease in the mortality rate, roughly equivalent to 2,300 patients, which represents a decrease of approximately 240 patients compared to the estimate obtained from considering only confounders.

It is crucial to note that, on average, the ATT scores are lower than the ATE and ATC scores. There could be other unquantifiable factors, such as socio-economic status and timely access to healthcare, that may contribute to patients who take PrEP being at a reduced risk for mortality. To mitigate this potential bias, a potential future direction could be to utilize instrumental variables as a means of eliminating such bias. [19].

## V. CONCLUSIONS

To the best of our knowledge, this is the first study to use causal modeling to measure the effects of PrEP on mortality in COVID-19-positive patients. We showed that taking PrEP results in a 2.1% decrease in the death rate, on average, in COVID-19 patients. The use of causal modeling can help us avoid the pitfall of controlling every observable variable, such as in “kitchen-sink” regression, and focus on observing only relevant variables like confounders, mediators, and colliders. Moreover, we can also compute counterfactuals to measure individual treatment effects for every patient and determine a personalized treatment policy. Our efforts included the integration of domain expertise in the generation of our causal graphs. In our future endeavors, we aim to investigate the use of instrumental variables and conduct sensitivity analysis in order to properly account for unmeasured confounding factors.

## Data Availability

All data produced in the present study are available upon reasonable request to the authors

